# HIV-1 RNA in Large and Small Plasmatic Extracellular Vesicles: a Novel Parameter for Monitoring Immune Activation and Virological Failure

**DOI:** 10.1101/2024.11.01.24316593

**Authors:** Julien Boucher, Wilfried Wenceslas Bazié, Benjamin Goyer, Michel Alary, Caroline Gilbert

## Abstract

**Background:** Antiretroviral therapy (ART) suppresses viral replication in most people living with HIV-1 (PLWH). However, PLWH remain at risk of viral rebound. HIV-1 infection modifies the content of extracellular vesicles (EVs). The changes in microRNA content in EVs are biomarkers of immune activation and viral replication in PLWH. Moreover, viral molecules are enclosed in EVs produced from infected cells. Our objective was to assess the value of EV-associated HIV-1 RNA as a biomarker of immune activation and viral replication in PLWH.

**Methods:** Plasma samples were obtained from a cohort of 53 PLWH with a detectable viremia. Large and small EVs were respectively purified by plasma centrifugation at 17,000 x *g* and by precipitation with ExoQuick™. HIV-1 RNA and microRNAs were quantified in the EV subtypes by RT-qPCR.

**Findings:** HIV-1 RNA content was higher in large EVs of ART-naive PLWH. Small EVs HIV-1 RNA was equivalent in ART-naive and ART-treated PLWH and positively correlated with CD4/CD8 T cell ratio. In ART-naive PLWH, HIV-1 RNA content of large EVs correlated with small EV-associated miR-29a, miR-146a and miR-155, biomarkers of viral replication and immune activation. A receiver operating characteristics analysis showed that HIV-1 RNA in large EVs discriminated PLWH with a high CD8 T cell count.

**Interpretation:** HIV-1 RNA in large EVs was associated with viral replication and immune activation biomarkers. Inversely, HIV-1 RNA in small EVs was related to immune restoration. Overall, these results suggest that HIV-1 RNA quantification in purified EVs could be a useful parameter to monitor HIV-1 infection.

**Funding:** Canadian Institutes of Health Research (CIHR) grants MOP-391232; MOP-188726; MOP-267056 (HIV/AIDS initiative)

**Research in context:** *Evidence before this study:* Antiretroviral therapy (ART) suppress viral replication to make HIV-1 infection manageable, but fails to clear the virus from people living with HIV-1 (PLWH). Hence, the infection becomes a chronic condition characterized by a dysfunction of the immune system caused by repeated activation and a persistent risk of a resurgence of viral replication (viral rebound). New biomarkers are required to improve the care of PLWH by identifying the individuals with a greater immune dysfunction and/or a higher risk of viral rebound. HIV-1 infection modifies the abundance, size and content of plasmatic extracellular vesicles (EVs). Specific host microRNAs enrcichment in EVs correlates with immune activation and viral rebound. In addition, viral proteins and genomic material are found within EVs. Various EV subtypes are released by infected cells, all using different biogenesis machinery. The distribution of HIV-1 RNA in EV subtypes has never been assessed and this novel parameter could provide information on the infection progression.

*Added value of this study:* This study provides the first quantification of HIV-1 RNA in two EV subtypes, large and small, from the plasma of PLWH. Large EVs HIV-1 RNA was lower in ART-treated PLWH and decreased with the duration of treatment. HIV-1 RNA associated to large EVs was a better predictor of immune activation than the standard plasma viral load. Inversely, the HIV-1 RNA concentration in small EVs was unaffected by ART and linked to better immune functions. Overall, the results presented in this study suggest that HIV-1 RNA in large EVs originates from ongoing viral replication, while HIV-1 in small EVs is the produce of proviral transcription.

*Implications of all the evidence:* The standard procedure for the clinical care of PLWH is to quantify HIV-1 RNA in the whole plasma, disregarding the context of its production. We show that the differential distribution of HIV-1 RNA in large and small EVs seems to be an indicator of disease progression. The purification of plasmatic EVs is considered as a non-invasive liquid biopsy to assess the progression of diseases. PLWH could benefit from the analysis of their plasmatic EVs to monitor the infection with an improved precision.

## 1. Introduction

Forty years have passed since the outbreak of the HIV-1 pandemic (1). The greatest progress in fighting HIV-1 infection was the discovery of molecules that inhibit HIV-1 replication (2). This led to the implementation of antiretroviral therapy (ART), which remains to this day the only way to manage HIV-1 infection (3). ART blocks HIV-1 replication in people living with HIV-1 (PLWH), but viral reservoirs persist. Consequently, ART must be taken daily by PLWH to avoid viral rebound and plasmatic viral load has to be measured every six months to ensure viremic control (4).

Virological failure is either caused by treatment resistance or loss of treatment adherence (5). Due to virological failure, CD4 T lymphocyte count plummets and the treatments must be optimized (6). Some PLWH have a detectable low-level viremia defined as non-suppressible viremia (NSV) (7, 8). Despite many years of ART, NSV occurs without the apparition of treatment resistance, and cannot be suppressed by treatment intensification (9). NSV originates from defective or replication-competent provirus transcriptional activity of infected T cell clones rather than new infection events (7). PLWH with NSV have an immune activation level similar to ART-suppressed PLWH (10). To summarize, a positive viral load assay could indicate an ongoing virus replication that requires immediate treatment optimization, or viral expression from latently infected cells for which treatment optimization is unnecessary (8). Thus, there is a need to understand better the mechanisms and the context behind detectable viremia in PLWH to facilitate their clinical care (8).

In recent years, the roles of extracellular vesicles (EVs) as biomarkers and a non-invasive tool to monitor the HIV-1 disease progression have emerged (11–13). EVs are nanoparticles enclosed by a bilayered lipid membrane (14). They are separated into two major subclasses depending on their origin. The first class, microvesicles, is named large EVs in this article. Large EVs are the product of the membrane budding (15). The second class is exosomes, called small EVs in this paper. Small EVs originate from the inward budding of endosomes and are released in the extracellular environment when endosomes merge with the plasma membrane(16). The endosomal sorting complex required for transport (ESCRT) machinery is the major pathway of small EVs biogenesis (16). Cargo selection in EVs subtypes is simultaneously a passive and active process. It is passive because cytoplasm and membrane content can be randomly selected in EVs. As a result, component concentration in EVs changes according to cellular expression. The EV biogenesis machinery drives the active cargo selection process. Consequently, active cargo selection varies between EV subtypes and can increase cargo concentration independently of cellular expression changes (17).

HIV-1 infection modifies the host microRNA content of EVs (18). Enrichment of miR-29a, miR-146a and miR-155 in small EVs of PLWH under ART was a predictor of detectable viremia (12). MiR-155 in large EVs of PLWH under ART was a predictor of immune activation (19). MiR-155-enriched EVs enhanced HIV-1 infection and promoted inflammation in recipient cells (18). Due to the similarities between EVs and HIV-1 biogenesis, infected cells release EVs harbouring viral RNA and proteins (20, 21). The unspliced genomic RNA of HIV-1 and the transactivation response element (TAR) RNA sequence were measured in the EVs released by infected cells (20–22). Inhibition of the ESCRT pathway lowers HIV-1 RNA concentration in EVs, showing its role in viral RNA sorting to EVs (22). Moreover, HIV-1 RNA in EVs persists despite ART, even when the plasma viral load is indetectable (23).

Since EV subtypes have different biogenesis pathways and we have shown differential biomarker roles for large and small plasmatic microRNA in EVs, we hypothesized that ART influences the viral RNA distribution in both EVs subtypes of PLWH. This new measurement could be a future biomarker for HIV infection management. The present study showed that large EVs were more abundant and contained more HIV-1 RNA in ART-naive PLWH and HIV-1 RNA in small EVs was unaffected by ART but correlated with the CD4/CD8 ratio.

## 2. Methods

### 2.1 Study participants

The cohort of PLWH present in this study (n = 53) was selected from a larger cohort of PLWH recruited in Bobo-Dioulasso and Ouagadougou (Burkina Faso) (12, 19, 24). These participants were selected because they had a detectable plasma viral load at their recruitment. All participants were anonymous volunteers and provided written informed consent.

### 2.2 Plasma EVs purification

Plasma EVs purification was performed according to a well-established procedure (11, 24, 25). Platelet-free plasma (250 µL) was thawed at room temperature and treated with proteinase K (1.25 mg/mL) for 10 minutes at 37 ℃ to eliminate protein aggregates and extravesicular RNA. Then, the plasma was centrifuged at 3,000 x *g* for 15 minutes to discard apoptotic vesicles, and at 17,000 × *g* for 30 minutes to pellet large EVs. The remaining supernatant was mixed with 63 µL ExoQuick™ (System Biosciences) reagent and incubated at 4 ℃ overnight. A centrifugation at 1,500 × *g* for 30 minutes precipitated small EVs. EV pellets were washed with 0.22 µm filtered PBS and resuspended in 250 µL of 0.22 µm filtered PBS.

### 2.3 Hydrodynamic size measurement

Quality, homogeneity and size of EV samples were analyzed by dynamic light scattering (DLS) with a Zetasizer Nano ZS (Malvern Instruments). Hydrodynamic diameter measurements were done in duplicate at room temperature.

### 2.4 EV quantification by flow cytometry

Absolute EVs quantification by flow cytometry was performed as previously described (12). EVs were stained with lipophilic carbocyanine DiD dye and CellTrace^TM^ CFSE (ThermoFisher Scientific), at a final concentration of 5 µM. DiD+ and CFSE+ events were considered EVs. EV concentration in our samples was determined with 15 µm silica beads (Polybead® Microspheres, Polysciences). The acquisition was performed on a modified BDFACS canto II with a photomultiplicator on the forward scatter (FSC) channel to improve nanoparticle detection.

### 2.5 RNA extraction

RNA was extracted from 50 µL of purified EVs (diluted in three volumes of Trizol® LS (ThermoFisher Scientific) using the phenol/chloroform method and resuspended in 15 µL of Tris/EDTA buffer (18, 26). RNA concentration was measured using a BioDrop™ spectrophotometer (Montreal Biotech Inc.).

### 2.6 HIV-1 RNA quantification

RT-PCR was performed on 5 µL of RNA with the Superscript IV reverse transcriptase kit (ThermoFisher Scientific) according to the manufacturer instructions with a final concentration of 10nM of primer set on a GeneAmp® PCR System 9700 (Applied Biosystems) (27). The primers were obtained from Integrated DNA Technologies: forward: 5’-GCCTCAATAAAGCTTGCCTTGA-3’; reverse: 5’-GGCGCCACTGCTAGAGATTTT –3′ to target the 3’ LTR region of the HIV-1 genome (NCBI accession number: K03455.1). CDNA was pre-amplified as described before (28). QPCR reactions were performed with the QuantiTech SyBr Green kit (Qiagen) according to the manufacturer’s instructions on a CFX384 Touch Real-Time PCR Detection System (Bio-Rad) (28).

### 2.7 MiRNA quantification by RT-qPCR

Reverse transcription was achieved using the miScript RT kit (Qiagen) in a GeneAmp® PCR System 9700 (Applied Biosystems) (18). Quantitative PCR was conducted in 96-well plates (Multiplate™, BioRad©) with miScript SyBr® Green from Qiagen© using a CFX Connect™ Real-time system (Bio-Rad©). The primers for miR-29a (Cat: MS00003262), miR-146a (Cat: MS00006566) and miR155 (Cat: MS00031489) were purchased from Qiagen©.

### 2.8 Statistical analysis

Normal and lognormal distribution tests were carried out for all data sets. Data sets with a lognormal distribution were transformed to obtain a normal distribution for statistical analysis. Statistical analyses were carried out using GraphPad Prism software version 10.2.2 with p-values below 0.05 considered statistically significant.

## 3. Results

### 3.1 HIV-1 RNA and microRNA distribution in large and small plasmatic EVs of PLWH

The distribution of viral RNA between large and small EVs and the impact of ART treatment were analyzed in a cohort of 53 PLWH with detectable viral load. Clinical characteristics presented in Table 1 show that 14 were ART-naive and 39 were ART-treated. The median age of ART-naive and ART-treated participants was 37 (interquartile range (IQR) 28-43) and 32 (27–43), respectively (p = 0.4165). ART-naive participants were living with HIV-1 for a median duration of 5 months (IQR 1-51) and ART-treated for 26 (IQR 10-43) (p = 0.2497). Treated participants were receiving ART for 24 months (IQR 7-66). The median CD4 T cell count, CD8 T cell count, and CD4/CD8 ratio were respectively 607 (IQR 264-762), 837 (IQR 515-1043) and 0.7 (IQR 0.4-1.2) for ART-naive participants and 332 (IQR 182-469), 667 (IQR 522-926) and 0.5 (IQR 0.2-0.7) for ART-treated participants. The plasma viral load median was higher for the ART-Naive at 18,622 copies/mL (IQR 4,472-55,063) than for the ART-treated participants at 9,373 (300–52,038).

**Table 1.** Characteristics of the study participants.

|  | ART-Naive<br>(n = 14) | ART<br>(n = 39) | *p*-value |
| --- | --- | --- | --- |
| Men, n (%) | 5 (35.7) | 19 (48.7) |  |
| Women, n (%) | 9 (64.3) | 20 (51.3) |  |
| Age (years); median (IQR) | 37 (28-43) | 32 (27-43) | 0.4165 |
| HIV duration (months);<br>median (IQR) | 5 (1-51) | 26 (10-43) | 0.2497 |
| CD4 T cells/μL; median<br>(IQR) | 607 (264-762) | 332 (182-469) | 0.0467 |
| CD8 T cells/μL; median<br>(IQR) | 837 (515-1043) | 667 (522-926) | 0.6006 |
| CD4/CD8 ratio; median<br>(IQR) | 0.7 (0.4-1.2) | 0.5 (0.2-0.7) | 0.0403 |
| ART duration (month);<br>median (IQR) | NA | 24 (7-66) |  |
| Plasma viral load<br>(copies/mL); median (IQR) | 18,622 (4,472-55,063) | 9,373 (300-52,038) | 0.6159 |

ART: antiretroviral therapy; IQR: Interquartile range; NA: not applicable

Large and small EVs were purified from the plasma of PLWH with a detectable viral load as previously described (11, 12). As expected, large EVs from ART-naive PLWH (324 ± 31 nm) and ART-treated (364 ± 24 nm) had a bigger hydrodynamic diameter than the small EVs of ART-naive PLWH (86 ± 8.9 nm) and ART-treated (87 ± 4.8 nm) (Figure 1A). Absolute quantification by flow cytometry revealed that ART-naive PLWH large EVs were more concentrated (1.52 x 10^7^ ± 1.80 x 10^6^ EVs/mL) than ART-treated PLWH (8.46 x 10^6^ ± 1.15 x 10^6^ EVs/mL) (Figure 1B). In addition, ART did not affect the count of small EVs since ART-naive PLWH small EVs were at a concentration of 1.76 x 10^7^ ± 3.51 x 10^6^ EVs/mL and ART-treated PLWH small EVs concentration was 1.35 x 10^7^ ± 2.67 x 10^6^ EVs/mL (Figure 1B).

**Figure 1.**
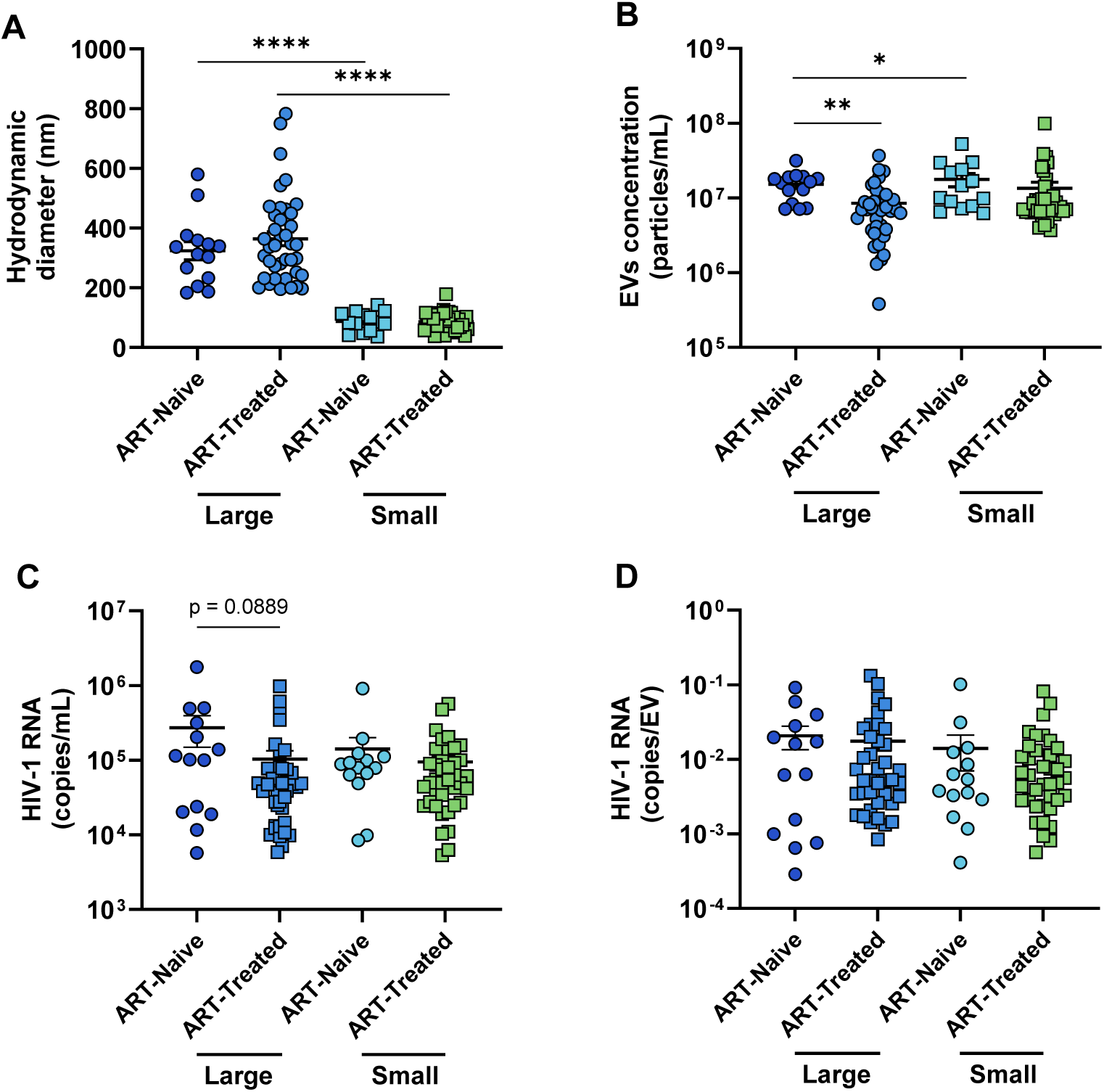
Viral RNA distribution in plasmatic EV subtypes from PLWH. EVs were purified from platelet-free and proteinase K-treated plasma by centrifugation at 17,000 x *g* to pellet large EVs, followed by ExoQuick precipitation to obtain small EVs. **A.** EV hydrodynamic size measurement by dynamic light scattering. **B.** Large and small EVs absolute concentration was determined by flow cytometry. **C.** Comparison of large and small EV-associated HIV-1 in ART-naive and ART-treated PLWH. **D.** The number of copies of HIV-1 RNA per mL of sample was divided by the number of EVs per mL of sample to estimate the number of HIV-1 copies per EV. Statistical analysis was carried out by one-way ANOVA (* p < 0.05; ** p < 0.01; **** p < 0.0001). ART: antiretroviral therapy; EV: extracellular vesicles; PLWH: people living with HIV-1.

Then, HIV-1 RNA was quantified by RT-qPCR in both types of EVs. HIV-1 RNA concentration was higher in large EVs (1.48 x 10^5^ ± 4.02 x 10^4^ copies/mL) and small EVs (1.07 x 10^5^ ± 2.09 x 10^4^ copies/mL) than in the plasma (5.57 x 10^4^ ± 1.61 x 10^4^ copies/mL) (Figure S1). The difference between ART-treated (2.72 x 10^5^ ± 1.24 x 10^4^ copies/mL) and ART-naive PLWH (1.03 x 10^5^ ± 3.05 x 10^4^ copies/mL) was most notable in large EVs (Figure 1C). HIV-1 RNA concentration in small EVs was similar in ART-naive (1.41 x 10^5^ ± 6.02 x 10^4^ copies/mL) and ART-treated PLWH (9.49 x 10^4^ ± 1.87 x 10^4^ copies/mL) (Figure 1C). With absolute EV concentrations, HIV-1 RNA copies per EV was calculated. In terms of HIV RNA per EV, differences between ART-naive (large: 0.021 ± 0.007 copies/mL; small: 0.014 ± 0.007 copies/mL) and ART-naive PLWH (large: 0.017 ± 0.005 copies/mL; small: 0.011 ± 0.003 copies/mL) were minimal (Figure 1D). Thus, the higher HIV-1 RNA content in large EVs of ART-naive PLWH was likely the result of increased production of large EVs containing HIV-1 RNA rather than the enrichment of HIV-1 RNA in large EVs. The results showed that ART decreased HIV RNA concentration in large EVs.

We previously showed a link between viral rebound and immune activation in PLWH and miR-29a, miR-146a and miR-155 content in plasmatic EVs (12, 19). Quantification of three microRNAs in large and small EVs by RT-qPCR showed that miR-155 was predominant in large EVs of ART-naive PLWH (Figure S2A-B), while miR-146 and miR-29a were significantly enriched in small EVs (Figure S2C-F). A correlation analysis showed that large EVs and small EVs HIV-1 RNA in ART-naïve PLWH correlated with miR-29a, miR-146 and to a lesser extent with miR-155 concentration in small EVs (Figure 2A and C). These results strengthened the viral rebound biomarker potential of miRNAs in small EVs. Conversely, in ART-treated PLWH, no correlation was found between large or small EVs HIV-1 RNA and small EVs miRNA content (Figure 2B and D). In addition, no correlation was found between large or small EVs HIV-1 RNA and large EVs miRNA content (Figure S3).

**Figure 2.**
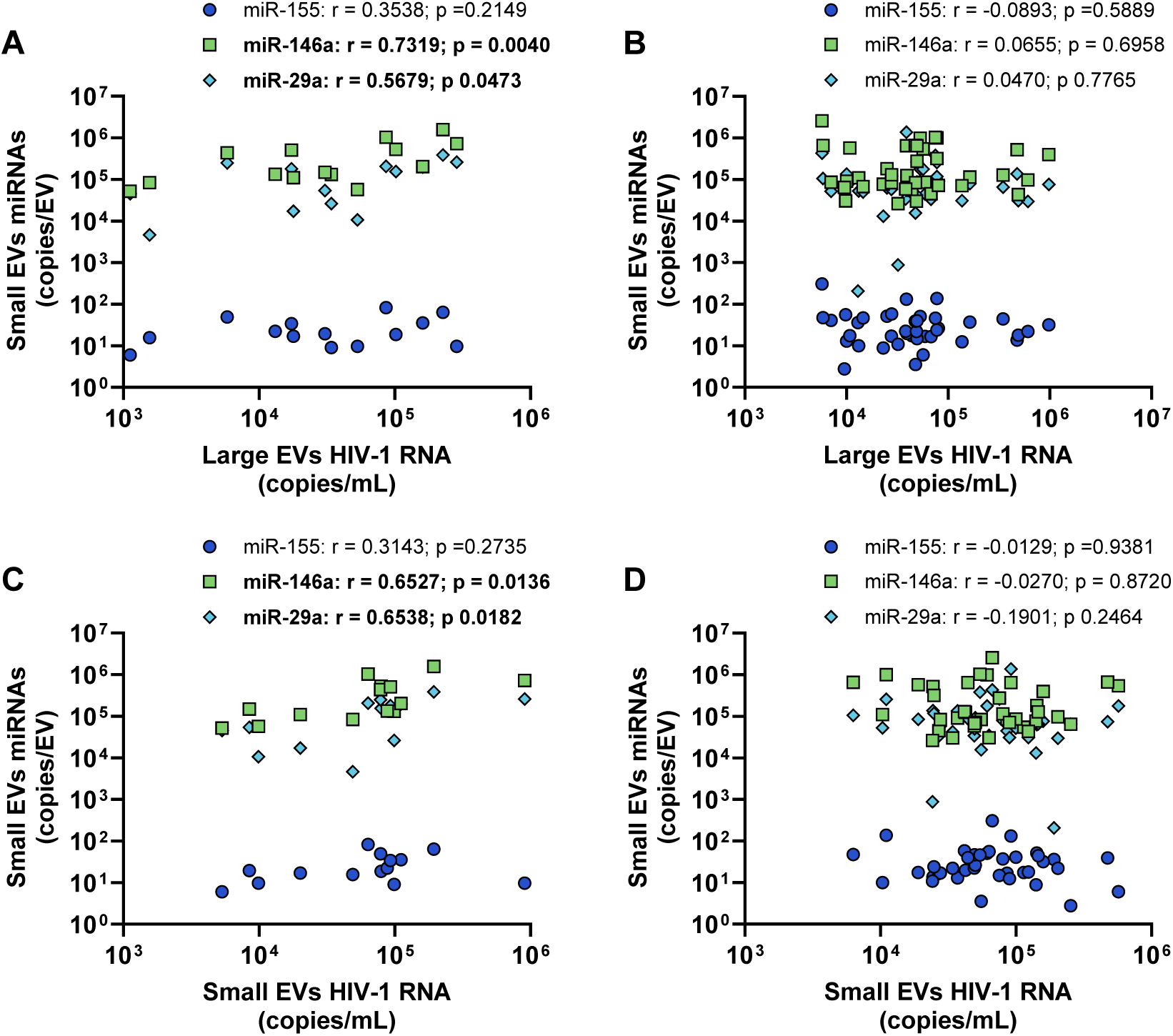
Separated correlation analysis between EV-associated viral load and miRNAs in ART-receiving and ART-naive PLWH. EVs were purified from platelet-free and proteinase K-treated plasma by centrifugation at 17,000 x *g* to pellet large EVs, followed by ExoQuick precipitation to obtain small EVs. Viral load and miRNAs content in EVs were quantified by RT-qPCR in large and small EVs. **A.** Correlation analysis between HIV-1 RNA concentration in large EVs and miRNAs concentration in small EVs of ART-naive PLWH. **B.** Correlation analysis between HIV-1 RNA concentration in large EVs and miRNAs concentration in small EVs of PLWH under ART. **C.** Correlation analysis between HIV-1 RNA concentration and miRNAs concentration in small EVs of ART-naive PLWH. **D.** Correlation analysis between HIV-1 RNA concentration and miRNAs concentration in small EVs of PLWH under ART. ART: antiretroviral therapy; EV: extracellular vesicles; PLWH: people living with HIV-1.

### 3.2 HIV-1 RNA in plasmatic EV subtypes of PLWH is associated with biomarkers of HIV-1 pathogenesis

Correlation analysis between large and small EVs HIV-1 RNA content and clinical parameters was performed to determine if HIV-1 RNA quantification in EVs is a better predictor of disease progression than the conventional plasma measurement. HIV-1 RNA measurement in the plasma correlated with the large EVs HIV-1 RNA measurement (Figure 3A). In ART-treated PLWH, large EVs HIV-1 RNA diminished with time under ART (Figure 3B). Conversly, in small EVs, HIV-1 RNA concentration was unaffected by ART (Figures 1C and 3B) There was a positive association between HIV-1 RNA in small EVs and the CD4/CD8 ratio (Figure 3C).

**Figure 3.**
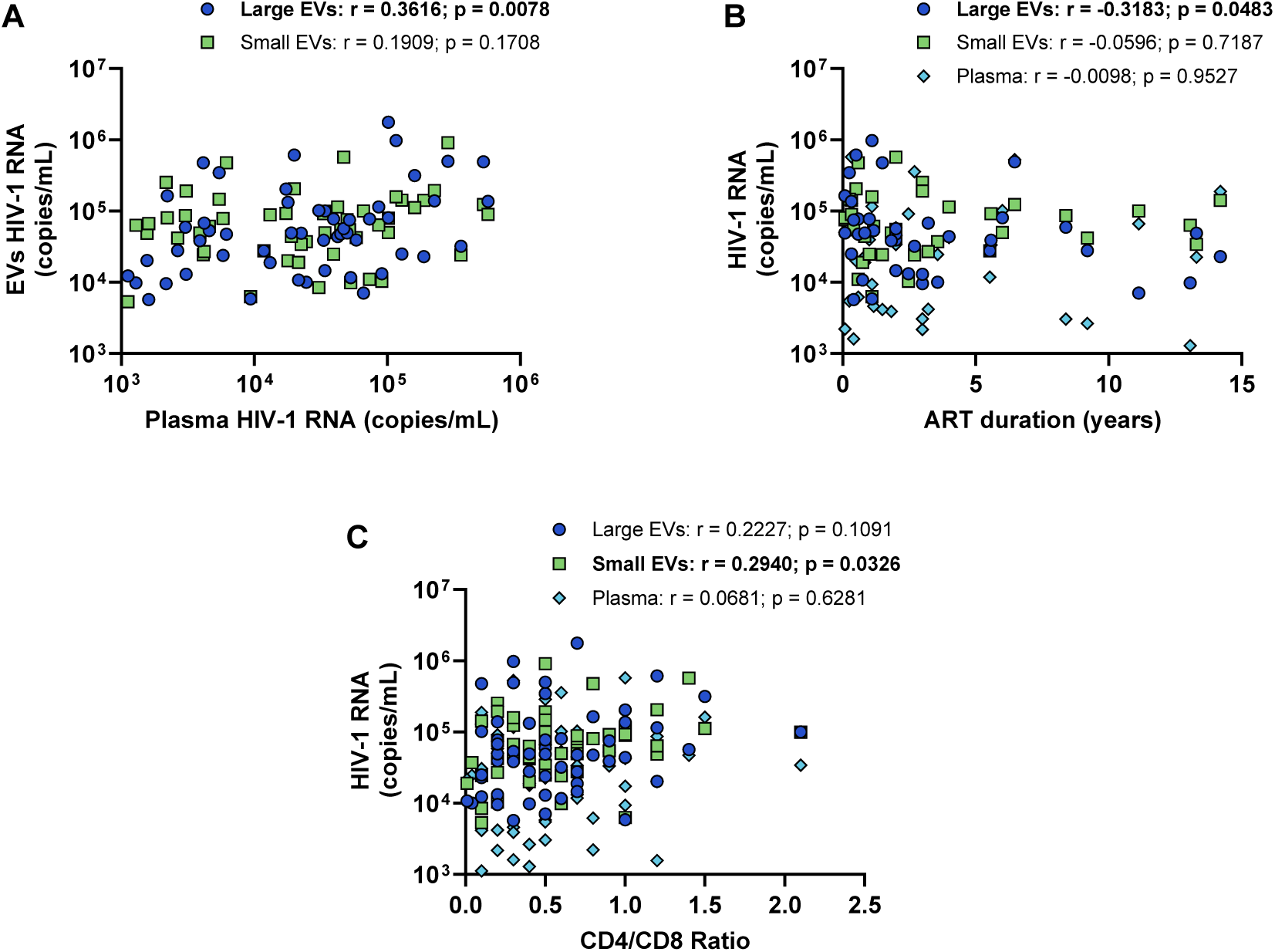
Correlation analysis between EV-associated HIV-1 RNA and biomarkers of infection progression. EVs were purified from platelet-free and proteinase K-treated plasma by centrifugation at 17,000 x *g* to pellet large EVs, followed by ExoQuick precipitation to obtain small EVs. HIV-1 RNA was quantified by RT-qPCR in large and small EVs. **A.** Correlation analysis between the plasma viral load and the purified EV-associated viral load. **B.** Correlation analysis between the time receiving ART and plasma and EVs HIV-1 RNA. **C.** Correlation between the CD4/CD8 ratio and plasma and EVs viral load. ART: antiretroviral therapy; EV: extracellular vesicles.

Next, we evaluated the potential of EV-associated HIV-1 RNA as a parameter to discriminate PLWH with immune activation and dysfunction. Among the 53 participants, those with a CD8 T cell count above 500 cells/µL were designated with immune activation (n = 41). The remaining participants with a CD8 T cell count below 500 cells/µL were in the control group (n = 12). A receiver operating characteristics (ROC) curve analysis was performed to determine which HIV-1 RNA concentration in the plasma, large EVs or small EVs discriminates PLWH with immune activation. HIV-1 RNA in the plasma (AUC = 0.50; 95% CI = 0.29-0.71; p = 0.9661) (Figure 4A) and the small EVs (AUC = 0.62; 95% CI = 0.43-0.80; p = 0.2257) (Figure 4C) could not distinguish PLWH with immune activation. HIV-1 RNA concentration in large EVs had the best diagnosis performance (AUC = 0.66; 95% CI = 0.50-0.83; p = 0.0852) (Figure 4B). PLWH with a CD4 count below 500 cells/µL (n = 38) or a CD4/CD8 ratio below 1 (n = 45) was designated with immune impairment. The participants with a CD4 T cell count above 500 cells/µL (n = 15) or a CD4/CD8 ratio above 1 (n = 8) were the controls. HIV-1 RNA in the plasma discriminated PLWH with immune impairment (Figure S4A and D), while HIV-1 RNA in large (Figure S4B-E) and small EVs (Figure S4C-F) did not. Overall, HIV-1 RNA in the plasma discriminated PLWH with low CD4 T cell count and CD4/CD8 ratio, while HIV-1 RNA in large EVs best discriminated PLWH with high CD8 T cell count.

**Figure 4.**
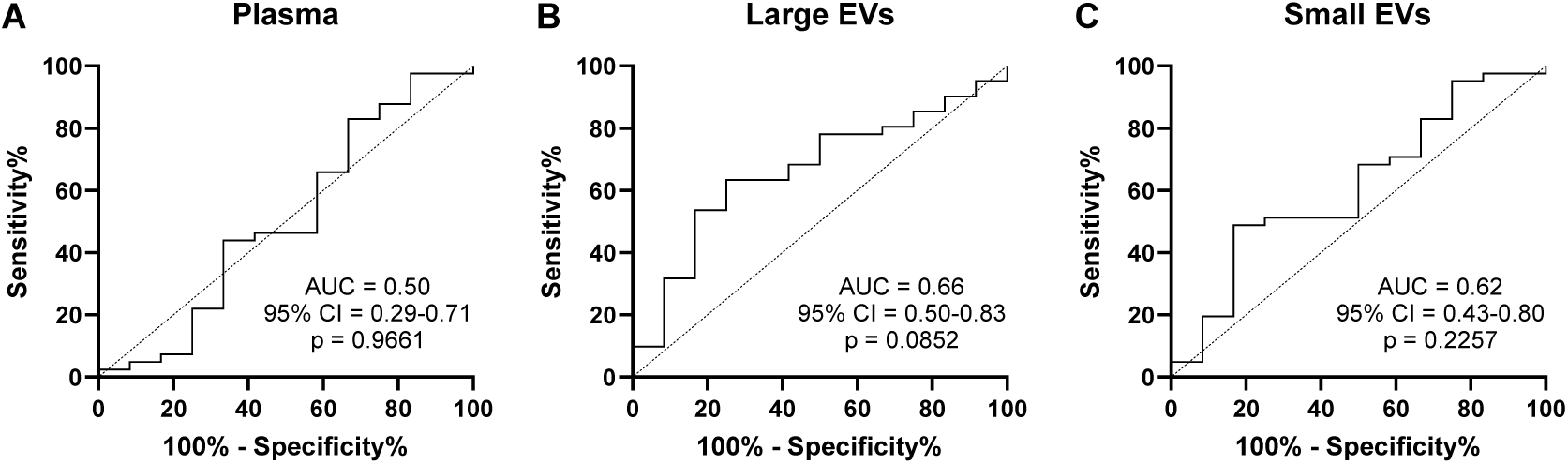
Diagnosis performance of immune activation by HIV-1 RNA concentrations in the plasma and EVs subtypes. HIV-1 RNA concentrations in the plasma and EV subtypes were used for ROC analysis to discriminate participants with immune activation. The participants with a CD8 T cell count below 500/µL were the controls. A CD8 T cell count above 500/µL defined immune activation. **A.** ROC curve of HIV-1 RNA concentration in the plasma. **B.** ROC curve of HIV-1 RNA concentration in large EVs. **C.** ROC curve of HIV-1 RNA concentration in small EVs. ROC: Receiver Operating Characteristics

## 4. Discussion

Decades of suppressive ART fails to clear viral reservoirs. Hence, PLWH are permanently at risk of viral rebound (4). When viremia is detected in PLWH, ART regimen modification or intensification is not always necessary and characterization of the virus in the plasma of PLWH could assist in HIV-1 infection management (8). This study aimed to quantify HIV-1 RNA in both types of plasmatic EVs of PLWH towards a more personalized HIV-1 infection management. We found that the value of small and large EV-associated HIV-1 RNA are biomarkers of disease progression and viral replication.

We previously reported an increase in total EVs concentration in ART-naive PLWH (13). Here, only the large EVs concentration was higher in ART-naive PLWH than PLWH under ART. Our results suggest that large EVs probably caused the increase in total EV concentration observed by Hubert et al. (13). The large EVs HIV-1 RNA level was higher in ART-naive PLWH than in treated PLWH. In PLWH under ART, HIV-1 RNA in large EVs was negatively correlated with time under ART. Interestingly, HIV-1 RNA concentration in small EVs was unaffected by ART. The persistence of HIV-1 RNA in small EVs despite ART is reminiscent of NSV, which is unaffected by ART (9). NSV has been more associated with men (29). Our results showed that men’s small EVs were richer in HIV-1 RNA than women’s (Figure S5). This data suggests again that the HIV-1 RNA in small EVs could be linked to NSV.

HIV-1 RNA in large EVs was susceptible to treatment, suggesting it is the product of active viral replication. These observations corroborate the previous assessment of viral RNA in large and small EVs in humanized mice (28). In that study, mice treatment lowered HIV-1 RNA in large EVs only. HIV-1 RNA in humanized mice small EVs correlated positively with the CD4 and CD4/CD8 ratio. The same correlation was observed in PLWH small EVs with a positive CD4/CD8 ratio. Viral replication causes a loss of CD4 T lymphocytes and expansion of CD8 T lymphocytes (6, 30). This strengthens the hypothesis that HIV-1 RNA in small EVs is not the product of viral replication. The hypothesis that large EVs contain HIV-1 RNA from active viral replication and small EVs contain HIV-1 RNA from provirus transcription could be explored by RNA sequencing. Viral replication will result in more genetic diversity due to the error-prone reverse transcriptase (31). On the contrary, HIV-1 RNA from NSV is less diverse since it comes from the transcriptional activity of clonally expanded CD4 T lymphocytes (7, 32). The Nanopore sequencing technology efficiently detects genetic divergence caused by viral replication (33) and mutations associated with ART resistance (34). Nanopore additionally offers the possibitiy to sequence the full length HIV-1 genome (35). This novel technology could further characterize the viral RNA associated with EVs subtypes.

This is the first report of comparative HIV-1 RNA quantification in EV subtypes of PLWH. Viral proteins and genetic material have been generally linked to small EVs (exosomes) because their biogenesis intertwines (36). Viruses can be formed in multivesicular bodies with exosomes (37). HIV-1 assembly also occurs at the plasma membrane (38). Both secretion mechanisms are involved in viral replication in infected cells but their relative contribution to HIV-1 pathogenesis has never been explored. Different biogenesis machinery incorporates disparate RNA and microRNA content in large and small EVs (39). This could explain the differential distribution of HIV-1 RNA in large and small EVs among PLWH. In addition, viruses would be associated with different host components whether they form at the plasma membrane or in multivesicular bodies. For example, miR-155 is enriched in large EVs during HIV-1 infection, promoting viral replication in the recipient cell (18). Therefore, viruses related to large EVs could be more infectious due to miR-155 transfer.

In this study, we showed a differential distribution of HIV-1 RNA in plasmatic EVs subtypes of PLWH. Viral replication was linked to HIV-1 RNA in large EV. Besides, HIV-1 RNA in small EVs was associated with immune restoration. This novel parameter could help us predict HIV-1 infection progression in PLWH and decipher the cause of a viral rebound. Combined with a miRNA analysis, we could establish a nucleic acid profile in EVs associated with immune dysfunction and virological failure to improve PLWH monitoring.

## Author contributions

Conceived and designed the experiments J.B., W.W.B., B.G. and C.G.: Performed the experiments: J.B., W.W.B. and B.G. Analyzed the data: J.B., W.W.B., B.G. and C.G.; Contributed clinical samples, reagents, materials, and analytical tools: J.B., W.W.B. M.A. and C.G.; Wrote the manuscript: J.B. and C.G. All authors have critically reviewed the paper and have agreed on the published version of the manuscript.

## Acknowledgements

This research was funded through Canadian Institutes of Health Research (CIHR) grants MOP-391232; MOP-188726; MOP-267056 (HIV/AIDS initiative) to C.G. J.B. and W.B.B. are the recipient of the Desjardins scholarship from the Fondation du CHU de Québec. J.B. and W.B.B. are recipients of the recruitment scholarship from the AIDS Research Fund of Université Laval. W.W.B. is the recipient of the leadership and sustainable development scholarship and the Fonds de Recherche du Québec – Santé (FRQ-S) doctoral training scholarship. The FRQ-S supports the Centre de recherche du CHU de Québec – Université Laval infrastructure. The authors thank Drs. Martin Pelletier and Stephane Gobeil for access to the qPCR platform. We are very grateful to the study participants, without whom this study would not have been feasible.

## Institutional review board statement

The study was conducted according to the guidelines of the Declaration of Helsinki and approved by the Centre de Recherche du CHU de Québec-Université Laval (Québec, QC, Canada) ethics review boards C12-03-208 (2012–2021) and CER-2019-4258. All subjects were volunteers and provided written informed consent before participating in the study.

## Informed consent statement

Written informed consent was obtained from all subjects involved in the study.

## Data availability statement

The study protocol, results and informed consent documents will be made available to researchers upon request from the corresponding author. Researchers will be asked to complete a concept sheet for their proposed analyses to be reviewed, and the investigators will consider the overlap of the proposed project with active or planned analyses and the appropriateness of the study data for the proposed analysis.

