## Supporting information for "HIV-1 RNA in Large and Small Plasmatic Extracellular Vesicles: a Novel Parameter for Monitoring Immune Activation and Virological Failure"

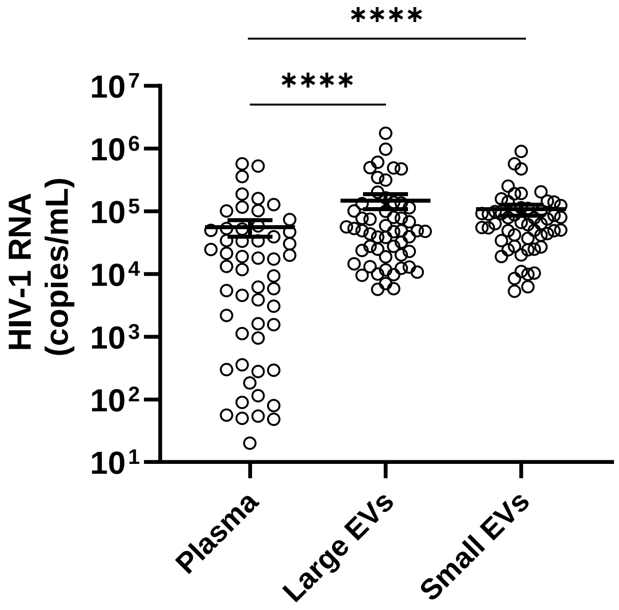


**Figure S1. Comparison of plasma viral load with EV-associated HIV-1 RNA in PLWH.**
EVs were purified from platelet-free and proteinase K-treated plasma by centrifugation at 17,000 x *g* to pellet large EVs, followed by ExoQuick precipitation to obtain small EVs. HIV-1 RNA was quantified in the total plasma and EVs of PLWH. A one-way ANOVA statistic test was performed (**** $p < 0.0001$). EV: extracellular vesicles; PLWH: people living with HIV-1.

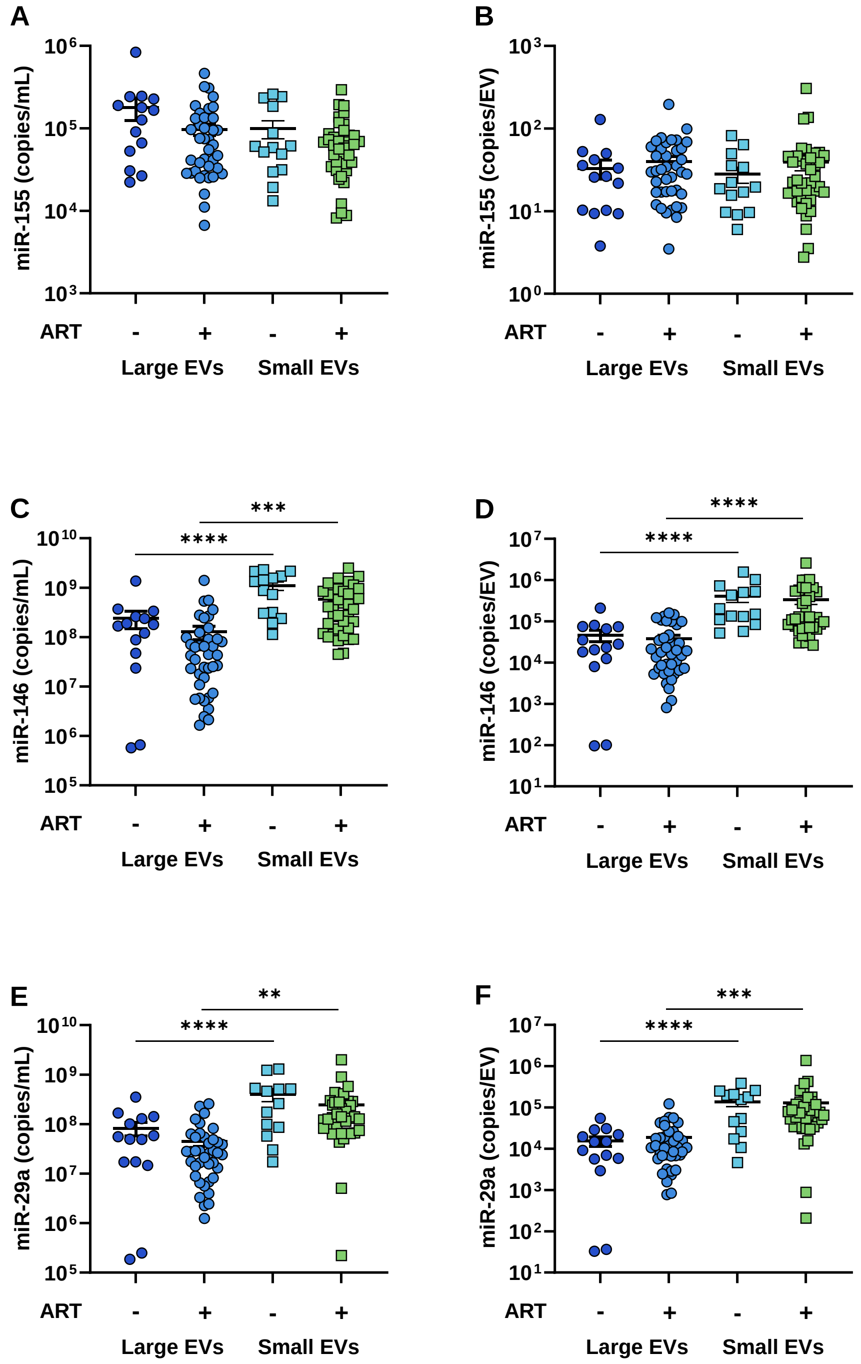


**Figure S2. Distribution of host miRNAs in large and small plasmatic EVs.**
EVs were purified from platelet-free and proteinase K-treated plasma by centrifugation at 17,000 x *g* to pellet large EVs, followed by ExoQuick precipitation to obtain small EVs. MiRNAs were quantified in EVs by RT-qPCR. The concentrations were presented as copies/mL. The number of copies per EV was calculated with the absolute EV counts obtained by flow cytometry. **A-B**. MiR-155. **C-D**. MiR-146. **E-F**. MiR-29a. Paired t-tests were performed for statistical analysis (** $p < 0.01$; *** $p < 0.001$; **** $p < 0.0001$). EV: extracellular vesicles.

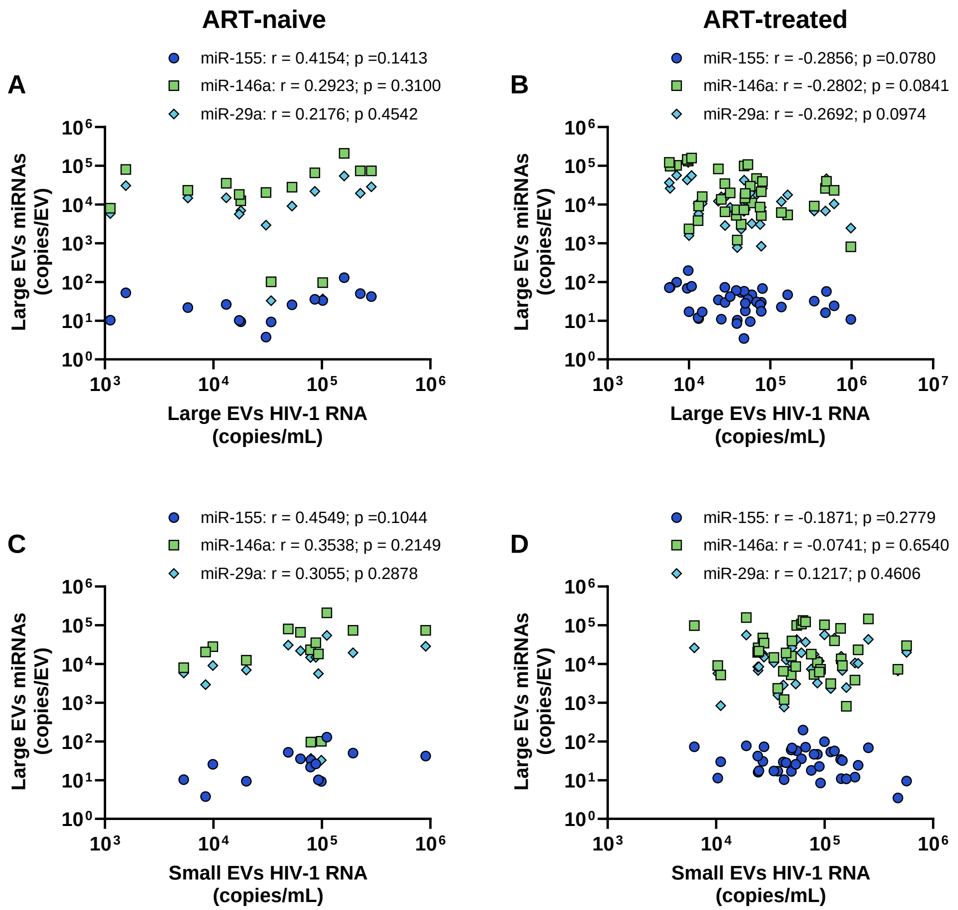


**Figure S3. Correlation analysis between EV-associated HIV- RNA and miRNAs in Large EVs from ART-treated and ART-naive PLWH.**

EVs were purified from platelet-free and proteinase K-treated plasma by centrifugation at 17,000 x *g* to pellet large EVs, followed by ExoQuick precipitation to obtain small EVs. HIV-1 RNA and miRNAs content in EVs were quantified by RT-qPCR in large and small EVs. **A.** Correlation analysis between HIV-1 RNA concentration in large EVs and miRNAs concentration in large EVs of ART-naive PLWH. **B.** Correlation analysis between HIV-1 RNA concentration in large EVs and miRNAs concentration in large EVs of ART-treated PLWH. **C.** Correlation analysis between HIV-1 RNA concentration in small EVs and miRNAs concentration in large EVs of ART-naive PLWH. **D.** Correlation analysis between HIV-1 RNA concentration in small EVs and miRNAs concentration in large EVs of ART-treated PLWH. ART: antiretroviral therapy; EV: extracellular vesicles; PLWH: people living with HIV-1.

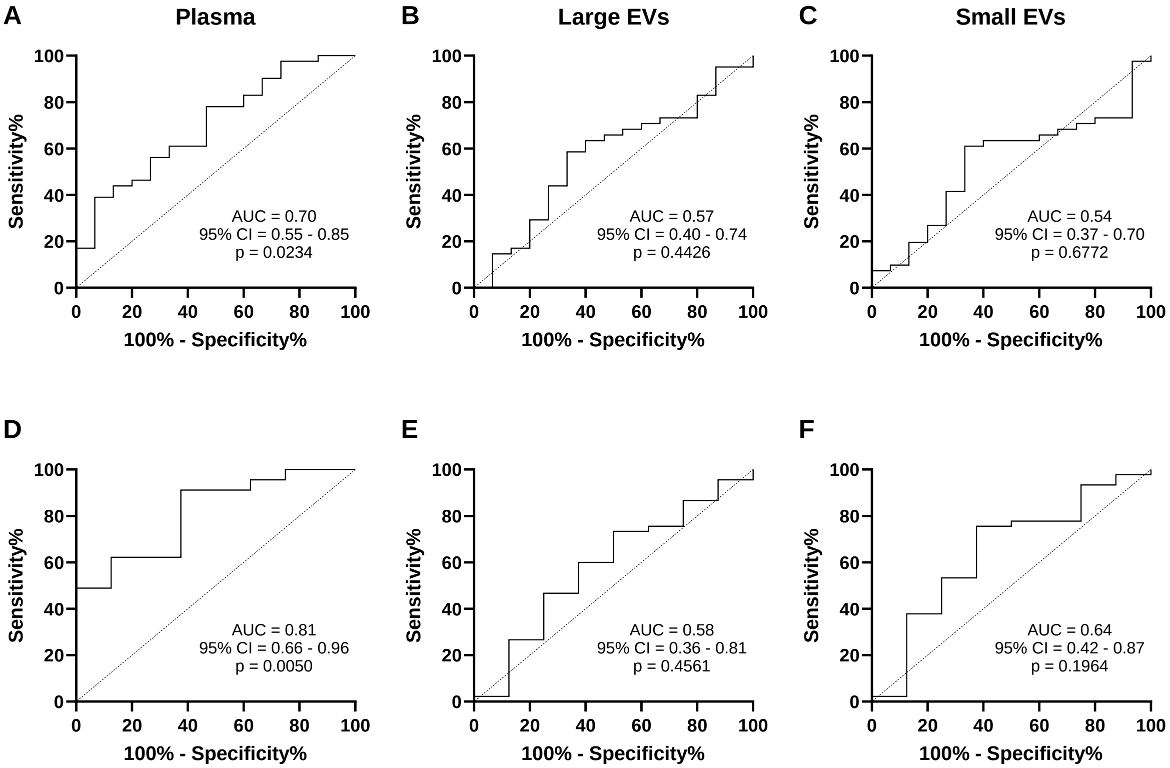


**Figure S4. Diagnosis performance of immune impairment by HIV-1 RNA concentrations in the plasma and EVs subtypes.**

HIV-1 RNA concentrations in the plasma and EV subtypes were used for ROC analysis to discriminate participants with immune impairment. For **A**, **B** and **C**, the participants with a CD4 T cell count above 500/µL were the controls (n = 15) and a CD4 T cell count below 500/µL defined immune impairment (n = 38). For **D**, **E** and **F**, the participants with a CD4/CD8 ratio above 1 were the controls (n =8) and a CD4/CD8 ratio below 1 defined immune impairment (n = 45). **A and D.** ROC curve of HIV-1 RNA concentration in the plasma. **B and E.** ROC curve of HIV-1 RNA concentration in large EVs. **C and F.** ROC curve of HIV-1 RNA concentration in small EVs. EV: extracellular vesicles; ROC: Receiver Operating Characteristics.

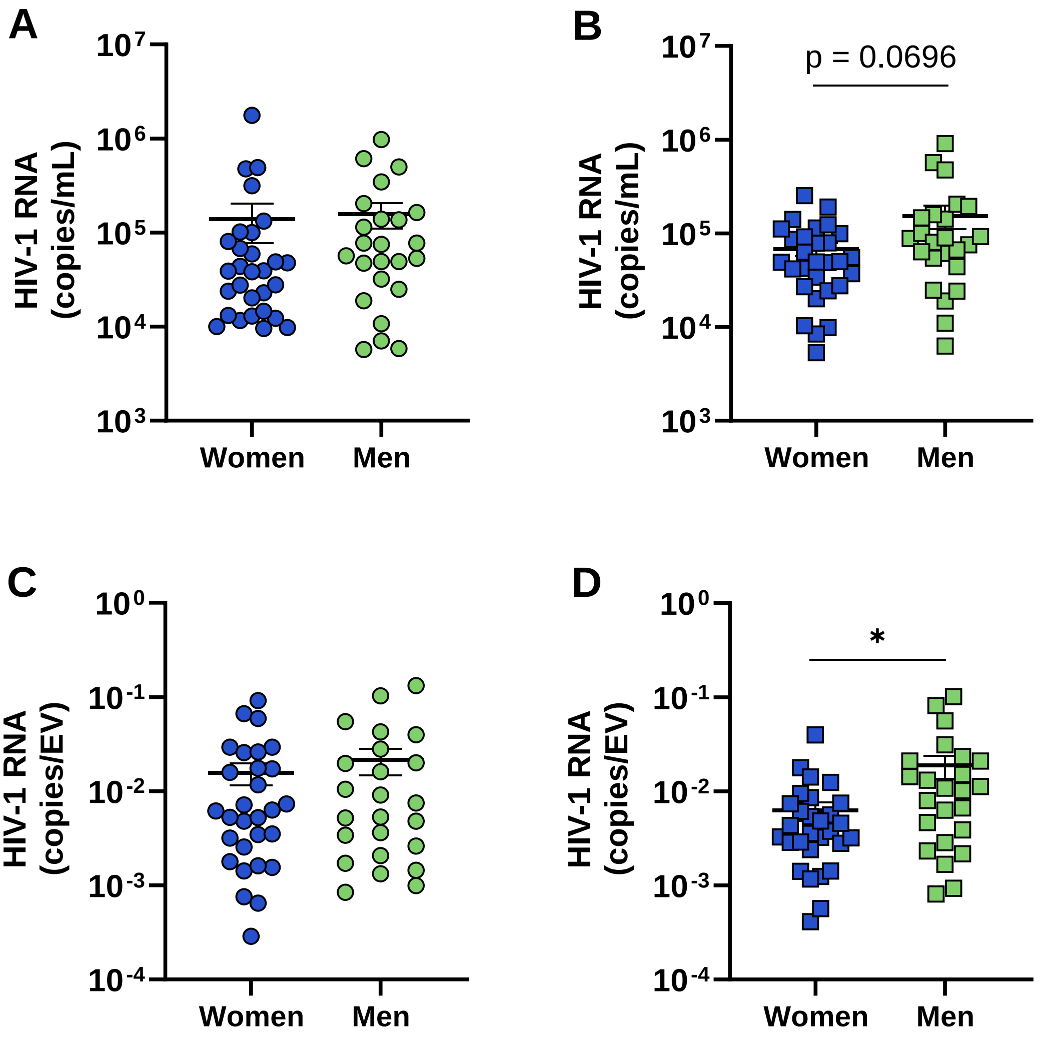


**Figure S5. Comparison of EV-associated HIV-1 RNA in women and men living with HIV-1.** EVs were purified from platelet-free and proteinase K-treated plasma by centrifugation at 17,000 x *g* to pellet large EVs, followed by ExoQuick precipitation to obtain small EVs. **A.** HIV-1 RNA in large EVs. **B.** HIV-1 RNA in small EVs. **C.** HIV-1 RNA in large EVs presented as copies per EV. **D.** HIV-1 in small EVs presented as copies per EV. An unpaired T-test was performed for statistical analysis (* < p 0.05). EV: extracellular vesicles.
